# The effect of respiratory activity, ventilatory therapy and facemasks on total aerosol emissions

**DOI:** 10.1101/2021.02.07.21251309

**Authors:** Nick M. Wilson, Guy B. Marks, Andrew Eckhardt, Alyssa Clarke, Francis Young, Frances L. Garden, Warren Stewart, Tim M. Cook, Euan R. Tovey

**Affiliations:** Department of Intensive Care Medicine, Prince of Wales Hospital, Sydney, Australia; Department of Anaesthesia, Royal Infirmary of Edinburgh, NHS Lothian, Scotland, UK; University of New South Wales, Sydney, Australia; Department of Intensive Care, Royal Prince Alfred Hospital, Camperdown, NSW, Australia; South Western Sydney Clinical School, University of New South Wales, Sydney, Australia; Ingham Institute of Applied Medical Research, Sydney, Australia; Department of Anaesthesia and Intensive Care Medicine, Royal United Hospital NHS Trust, Bath, UK; Bristol Medical School, University of Bristol, UK; Woolcock Institute of Medical Research, University of Sydney, Glebe, NSW, Australia

## Abstract

**Background:** Exhaled respirable aerosols (<5 µm diameter) present a high risk of severe acute respiratory syndrome coronavirus-2 (SARS-CoV-2) transmission. Many guidelines recommend using aerosol precautions during ‘aerosol generating procedures’ (AGPs) and droplet (>5 µm) precautions at other times. However, there is emerging evidence that respiratory activities such as cough and not AGPs are the important source of aerosols.

**Methods:** We used a novel chamber with an optical particle counter sampling at 100 L/min to count and size-fractionate all exhaled particles (0.5-25 µm). We compared emissions from ten healthy subjects during respiratory ‘activities’ (quiet breathing, talking, shouting, forced expiratory maneuvers, exercise and coughing) with respiratory ‘therapies’ designated as AGPs: high flow nasal oxygen (HFNO) and single or dual circuit non-invasive positive pressure ventilation, NIPPV-S and NIPPV-D, respectively. Activities were repeated wearing facemasks.

**Results:** Compared to quiet breathing, respiratory activities increased particle counts between 34.6-fold (95% confidence interval [CI], 15.2 to 79.1) during talking, to 370.8-fold (95% CI, 162.3 to 847.1) during coughing (p<0.001). During quiet breathing, HFNO at 60 L/min increased counts 2.3-fold (95% CI, 1.2 to 4.4) (p=0.03) and NIPPV-S and NIPPV-D at 25/10 cm H_2_O increased counts by 2.6-fold (95% CI, 1.7 to 4.1) and 7.8-fold (95% CI, 4.4 to 13.6) respectively (p<0.001). During activities, respiratory therapies and facemasks reduced emissions compared to activities alone.

**Conclusion:** Talking, exertional breathing and coughing generate substantially more aerosols than the respiratory therapies HFNO and NIPPV which can reduce total emissions. The risk of aerosol exposure is underappreciated and warrants widespread targeted interventions.

## Introduction

Severe acute respiratory syndrome coronavirus-2 (SARS-CoV-2) infection and consequent coronavirus disease-19 (COVID-19) is a significant cause of mortality and morbidity amongst patients, healthcare workers and the general population.^1,2^ Many international COVID-19 guidelines state that SARS-CoV-2 transmission is primarily through larger respiratory fluid ‘droplets’ (>5 µm diameter), while aerosols (<5 µm) are only a significant risk during ‘aerosol generating procedures’ (AGPs).^3,4^ Therefore, standard protection against COVID-19 is based on preventing droplet transmission, which includes surgical facemasks, whereas, fit-tested N95-rated respirators and enhanced environmental ventilation are recommended during AGPs.^3,4^ Aerosols are of concern as they can contain replication-competent virus, travel on airflows, better evade surgical masks and deposit on the alveolar epithelium, potentially increasing disease severity.^5–9^ Concerningly, a higher prevalence of infection has been observed in healthcare workers caring for COVID-19 patients using droplet compared with aerosol measures.^10–12^

The special status accorded to AGPs is based on weak epidemiological evidence from the SARS-CoV-1 epidemic where increased disease transmission occurred in healthcare workers exposed to patients requiring acute respiratory therapies.^13^ Aerosols were not measured in these studies.^13^ The respiratory therapies high flow nasal canula therapy (HFNO) and non-invasive positive pressure ventilation (NIPPV) are universally designated AGPs.^14^ However, these therapies may supress aerosol emissions by altering pulmonary mechanics or filtering exhaled gases.^15^ Earlier studies quantifying aerosols during therapies suggest both increased and decreased emissions.^16–18^ A recent study and pre-publication suggest coughing may generate up to 3-10 times more aerosols than HFNO and NIPPV.^19,20^ However, the methods used in these studies may have underestimated total emissions and exposure risk.^19,20^

Misclassification of HFNO and NIPPV as AGPs may have two serious adverse consequences. First, that the risk from common respiratory activities is underestimated so effective precautions are not widely used and second, patients may have delayed or restricted access to beneficial therapies.^3,14,21^

Based on the established mechanisms of physiological aerosol generation we hypothesised total emissions will be increased by exertional respiratory activity and decreased by clinically indicated therapies.^9,15^ To provide better quantification of risk, we developed a novel chamber to measure total human aerosol emissions during six respiratory activities, and compared them to emissions using HFNO, NIPPV and the wearing of surgical facemasks.

## Methods

We recruited healthy, non-smoking, healthcare workers, using a screening questionnaire and physiological measurements. The protocol was approved by the South Eastern Sydney Ethics Committee (ETH01467/2020) and written consent obtained.

The chamber was designed using clean airflow concepts from Morawska’s expiratory droplet investigation system (EDIS) and a large sampling cone from Milton’s Gesundheitt-II (Figure 1).^22,23^ The cone was connected to an optical particle counter (OPC) sampling at 100L/min (Aerotrak 9500, TSI Instruments, Minnesota, USA). The OPC counts particles into six size categories (‘bins’), (0.5-0.7, 0.7-1, 1-3, 3-5, 5-10, 10-25 µm).

**Figure 1.**
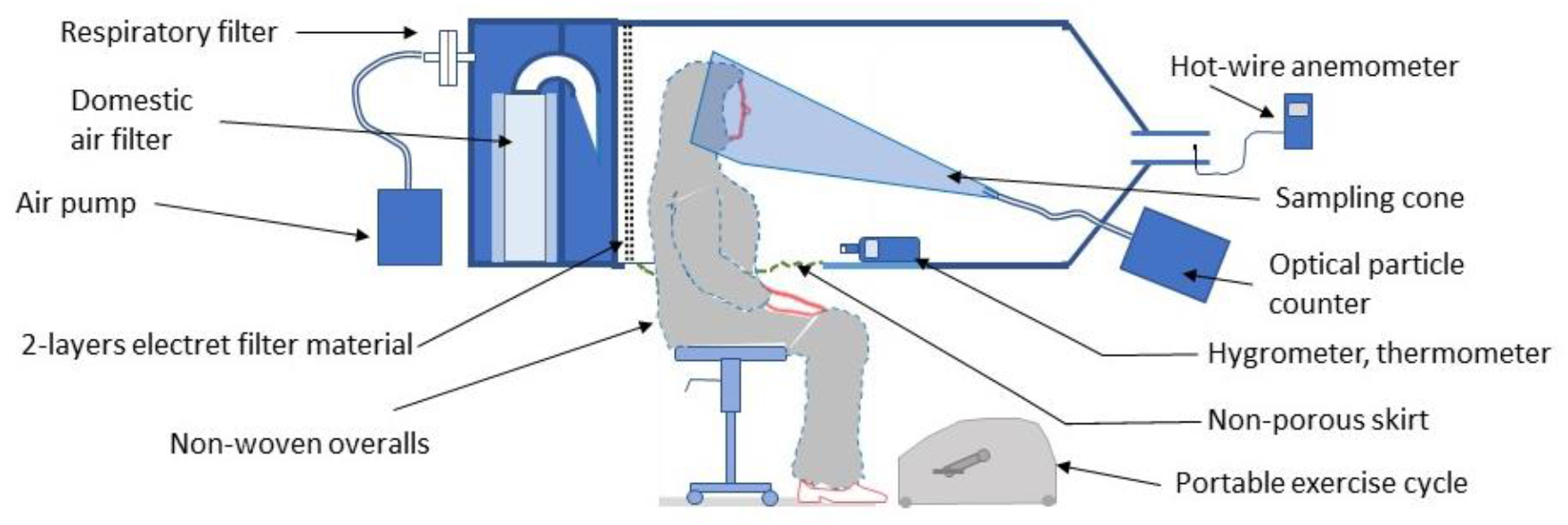
The sampling chamber consisted of a rear section, containing filters, which supplied clean air through a wall composed of air-filter media and a clear-walled forward section accommodated the torso of the subject. A flexible non-porous skirt enabled entry of the subject and the tubing of non-invasive devices. The subject’s head was positioned within a cut-away section of a large cone, which was attached to an optical particle counter, sampling at 100 L/min and mounted outside the chamber. Airflow in the 100 mm diameter tube at the distal end in the chamber was monitored via the anemometer probe. Humidity and temperature were monitored using a hygrometer and thermometer positioned in front of the subject, on the chamber floor. A moveable pedal exerciser was mounted so the subject could exercise in their seated position. A detailed description is provided in the Supplementary material.

Subjects wore hooded polypropylene coveralls and were positioned with their heads within the cone. During quiet breathing the chamber was purged, whereby counts fell from ambient (∼50,000 -70,000 / 100 L) to <120 total counts/100 L, (0.0012 particles/cc) and were stable (change <2 counts/second). Each sample required a one-minute prior purge, followed by one-minute of activity and sampling, and ended with a sustained purge. Exercise was an exception, where pedalling began one minute before sampling. The entire protocol lasted approximately four hours per subject. Six subjects performed the protocol in the order described and four in the reverse order.

Ten subjects performed six respiratory activities with and without surgical facemasks and then repeated selected activities while receiving three respiratory therapies designated as AGPs detailed in the study design (Supplementary Table S1). The six respiratory activities were chosen because they represent common aerosol generating activities or were proxies for respiratory symptoms associated with acute infections, such as increased work of breathing and atelectasis. The combinations of activities and therapies were based on practical feasibility and clinical relevance. The activities were:

- Quiet breathing through either the nose or mouth.
- Talking; repeating the alphabet while talking loudly.
- Shouting; repeating a short sentence as loud as could be sustained for this time.
- Six forced expiratory volume (FEV) manoeuvres – a full inspiration, and then full exhalation to residual volume. Prompted at 7-8-second intervals, finishing at 45 seconds.
- Six volitional coughs -moderate intensity, with timings as for FEVs.
- Exercise with a pedal exerciser (PhysioRoom, Padiham, United Kingdom) set to mid-load (to achieve ∼70% of maximal estimated heart rate).

The six activities were repeated wearing a surgical mask with ties – (Med-Con, item 170515, Shepparton, Australia).

The three respiratory therapies were as follows:

- A C6 ventilator (Hamilton, Bonaduz, Switzerland) delivered humidified 33°C HFNO at an FiO_2_ of 0.25, via a Optiflow plus circuit and MR850 humidifier (Fisher and Paykel, Auckland, Aotearoa) Flows were delivered at 20, 40 and 60L/minute during quiet breathing, and at 60 L/minute during talking, FEV, coughing and exercise.
- A V60 ventilator (Phillips, Eindhoven, Holland) delivered humidified 33°C NIPPV at an FiO_2_ of 0.25 via a Nivairo facemask (RT045, Fisher and Paykel), using an open single limb circuit with open expiratory port positioned inside the sampling cone. Pressures delivered were Inspiratory peak/expiratory peak airway pressures (IPAP/EPAP cm H_2_0) of 5/5, 10/10, 15/10, 20/10 and 25/10 and during exercise at 20/10 cm H_2_0.
- A Hamilton-C6 ventilator was used to deliver NIPPV-D using the same pressure ranges, facemask and using a dual limb circuit with a high-efficiency air particulate (HEPA) filter on the expiratory limb.

### Analysis

Sample size was based on pilot data obtained during the protocol development and prior studies.^24^ As particle counts were positively skewed and included some zero values, the counts were log transformed with a zero offset, as follows: log-count=log_10_ (count+0.3). To compare differences in particle counts between activities and respiratory therapies, a mixed effects linear regression model was used to take account of repeated measures across particle bin sizes and within participants (in Proc Mixed, SAS version 9.4). In all models the dependent variable was log-count. The main fixed effect was activity or therapy (or both). Reference values were quiet breathing (for activity) and no therapy. To assess the impact of wearing a facemask the models were repeated with the main fixed effect terms of activity, with and without a facemask. Results are reported as fold differences (95% confidence intervals) between the geometric means of activities and/or therapy. Particle counts in the six size bins were transformed to estimated particle volumes using the formula, volume = 4/3*π*radius^3^ (Table S2). Further details of the data analysis are provided in the appendix.

### Post Hoc Experiments

Subjects’ perception of air passing backwards during certain activities, prompted an observational study of the behaviour of visible expired plumes using e-cigarette aerosols and a replica transparent cone and two subjects. An unexpected difference in aerosol emissions between NIPPV-S and NIPPV-D prompted a detailed comparison of ventilator performance in two subjects. Both experiments are detailed in the supplementary methods.

## Results

### Subject

Four females and six males, mean age 29 years (SD=2.8), were recruited. Detailed subject and environmental data are in the appendix (Table S3, S4). We sampled 31,000 litres of air to make 1860 measurements (186/subject) of particle size and number.

### Respiratory activities

When compared to quiet breathing, each of the respiratory activities was associated with a large increase in emissions (Figure 2, Table 1, Table S6). This increase ranged from 34.6-fold during talking up to 370.8-fold during six coughs (all p<0.001). The fold increases, also presented as average total number of particles per activity and as estimated total volumes (Table S6).

**Table 1.** Overall fold change in average total particle counts relative to reference activity -quiet breathing or the same activity without the respiratory therapy or surgical facemask. Abbreviations: HFNO, high flow nasal canula therapy with flow in L/min; NIPPV-S and NIPPV-D, non-invasive positive pressure ventilation (single and dual circuits, respectively) with inspiratory/expiratory airway pressures shown in cm H_2_O.

| **Fold changes of average total particle counts during respiratory activities compared to quiet breathing alone.** |  |  |  |
| --- | --- | --- | --- |
| **Respiratory activity** | **Fold change** | **95% confidence interval** | **P value** |
| Talking | 34.6 | 15.2, 79.1 | <0.001 |
| Exercise | 58.0 | 25.4, 132.5 | <0.001 |
| Shouting | 163.6 | 71.6, 373.9 | <0.001 |
| Forced expirations | 227.6 | 99.6, 520.0 | <0.001 |
| Coughing | 370.8 | 162.3, 847.1 | <0.001 |
| **Fold changes of average total particle counts during respiratory therapies compared to quiet breathing alone.** |  |  |  |

| Therapy | Flow | Fold change | 95% confidence interval | P value |
| --- | --- | --- | --- | --- |
| HFNO | 20 | 1.3 | 0.6, 2.4 | 0.47 |
|  | 40 | 1.7 | 0.9, 3.3 | 0.10 |
|  | 60 | 2.3 | 1.2, 4.4 | 0.03 |
| Therapy | Pressure |  |  |  |
| NIPPV-S | 5/5 | 1.5 | 0.9, 2.3 | 0.08 |
|  | 10/10 | 1.3 | 0.9, 2.1 | 0.19 |
|  | 15/10 | 2.1 | 1.4, 3.3 | <0.001 |
|  | 20/10 | 2.4 | 1.5, 3.8 | <0.001 |
|  | 25/10 | 2.6 | 1.7, 4.1 | <0.001 |
| NIPPV-D | 5/5 | 1.9 | 1.1, 3.3 | 0.03 |
|  | 10/10 | 2.9 | 1.6, 5.0 | <0.001 |
|  | 15/10 | 3.1 | 1.7, 5.4 | <0.001 |
|  | 20/10 | 4.6 | 2.6, 8.0 | <0.001 |
|  | 25/10 | 7.8 | 4.4, 13.6 | <0.001 |
| **Fold changes of average total particle counts during respiratory therapies and exercise compared to exercise alone.** |  |  |  |  |
| Therapy |  | Fold change | 95% confidence interval | P value |
| HFNO 60 |  | 0.7 | 0.4, 1.3 | 0.22 |
| NIPPV-S 20/10 |  | 0.4 | 0.2, 0.7 | 0.02 |
| NIPPV-D 20/10 |  | 0.7 | 0.4,1.3 | 0.21 |
| **Fold changes of average total particle counts with HFNO 60L/min during activities compared to the same activities alone.** |  |  |  |  |
| Respiratory Activity |  | Fold change | 95% confidence interval | P value |
| Quiet breathing |  | 2.3 | 1.1, 4.9 | 0.03 |
| Talking |  | 0.9 | 1.0, 1.5 | 0.73 |
| Forced expirations |  | 0.7 | 0.4, 1.3 | 0.19 |
| Coughing |  | 0.5 | 0.3, 1.0 | 0.03 |

**Table 1** – Overall fold change in average total particle counts relative to reference activity - quiet breathing or the same activity without the respiratory therapy or surgical facemask.
| Fold changes of average total particle counts whilst wearing a surgical facemask during activities compared to without a facemask. |  |  |  |
| --- | --- | --- | --- |
| Respiratory Activity | Fold change | 95% confidence interval | P value |
| Overall | 0.4 | 0.3, 0.6 | <0.001 |
| Quiet breathing | 1.2 | 0.6, 2.2 | 0.62 |
| Talking | 0.4 | 0.2, 0.9 | 0.03 |
| Exercise | 0.7 | 0.4, 1.1 | 0.08 |
| Shouting | 0.3 | 0.1, 0.6 | 0.01 |
| Forced expirations | 0.3 | 0.1, 0.8 | 0.02 |
| Coughing | 0.2 | 0.1, 0.6 | 0.01 |

**Figure 2.**
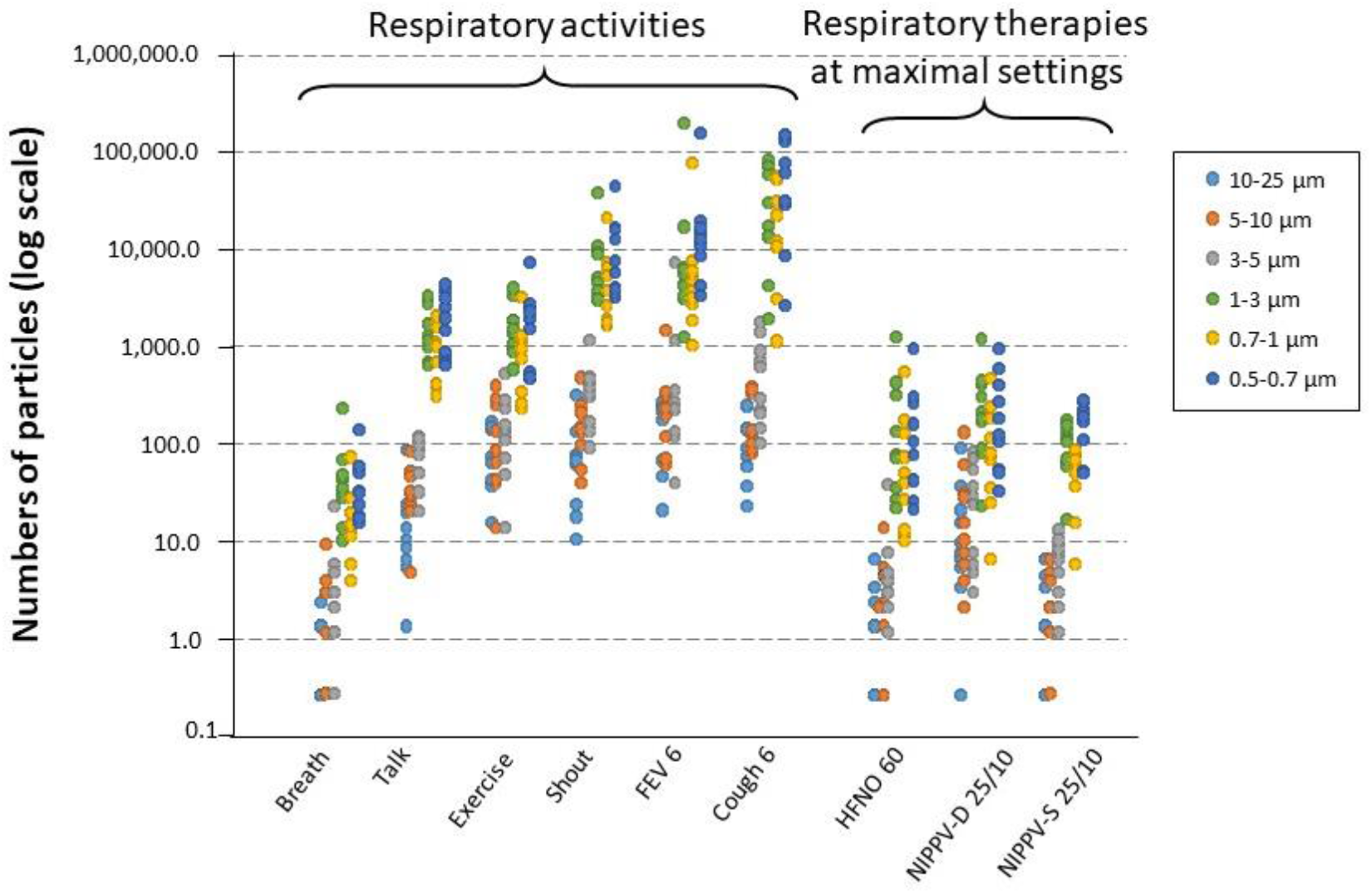
Shows the total numbers of exhaled respiratory particles sampled from ten subjects. Samples were measured over a period of one minute during six respiratory activities and when breathing quietly while three respiratory therapies, designated as ‘aerosol generating procedures’ (AGPs), were applied. The therapies are as follows: High flow nasal canula therapy (HFNO) and single or dual circuit non-invasive positive pressure ventilation, NIPPV-S and NIPPV-D, respectively. All respiratory therapies shown were recorded at the highest settings used: HFNO at a flow of 60 L/min and both NIPPV-S and NIPPV-D at inspiratory/expiratory airway pressures of 25/10 cm H_2_O. The size range in the six particle bins as measured by the optical particle counter are shown in the key. A value of 0.3 was added to all counts, so zero particle counts are shown as 0.3. Overlapping dot points are not shown. FEV – force expiratory volume maneuver. Both FEV and cough were repeated six times in the sampling minute.

### Respiratory therapies

The three respiratory therapies showed slight increases in total particle counts at higher flows and pressures (Table 1, S6, Figure 2) relative to the breathing benchmark. During HFNO the only significant increase occurred at a flow of 60 L/min where counts approximately doubled (p=0.03). During NIPPV-S, counts increased 2-3-fold, reaching significance at 15/10 cm H_2_O (p<0.001). With NIPPV-D, counts increased 2-8-fold and were significant at all pressures (all p≤0.03).

Particle counts reduced when HFNO was used during respiratory activities, significantly during coughing, where emissions were halved (p=0.03). During exercise, the three respiratory therapies reduced particle counts by 30-60%, though only significantly during NIPPV-S (p=0.02) (Table 1).

### Surgical facemasks

The effect of surgical facemasks varied with activity, generally decreasing total emissions with apparent larger reductions observed in activities with higher particle counts. Forced expiration and cough demonstrated approximately 3- and 5-fold reductions, p=0.02 and p=0.01 respectively (Table 1). The 1.2-fold increase in emissions wearing a mask during quiet breathing was not significant (p=0.62).

### Distribution of aerosols between activities and procedures

The overwhelming majority (>92%) of particles produced across all activities and procedures were respirable aerosols (≤5 µm) (Table S6). The proportion of the total volume of particles that was aerosols, ranged between 5.9% and 34.9% for all respiratory activities alone, with coughing producing the greatest proportion, and between 7.1 and 22.4% during HFNO and NIPPV with quiet breathing.

### Inter-subject variation

The intraclass correlation of subjects for all activities with and without devices was 0.065 and 0.068, respectively, indicating substantial variation between subjects in the total number of exhaled particles and the effect of activities and respiratory therapies. Although breathing was consistently a minor contributor to the total volume of particles, the ranking of other activities varied between subjects. The removal of the highest contributor had negligible effect on the relative magnitude of changes in particle emissions between activities and therapies (Figure S2, Table S8).

### Post Hoc Experiments

The qualitative visualisation of exhaled e-cigarette aerosols suggests minor incomplete sampling may have occurred with all activities, and substantial under-sampling during coughing, FEV and wearing of surgical masks (Video S1). The assessment of ventilator performance demonstrated NIPPV-D was associated with 30% more asynchronous respiratory cycles compared to NIPPV-S (Table S7).

## Discussion

This study is the first to explore (close to) complete exhaled respiratory emissions and the most detailed in comparing emissions (counts, size distributions and estimated volumes) across a broad range of respiratory activities with three non-invasive respiratory therapies. We demonstrate that the total emissions per minute during common respiratory activities are often one to two orders of magnitude greater than during HFNO and NIPPV which are currently classified as AGPs. Importantly, when these therapies were used during respiratory activities, emissions were reduced compared to the activities alone.

Our study advances and compliments two recent important studies which also compared respiratory activities and therapies.^19,20^ Gaeckle found NIPPV and HFNO did not generate significantly more aerosols compared to other respiratory activities, while coughing increased emissions 3-fold.^19^ Hamilton reports coughing produces a ten-fold rise in emissions, NIPPV-S was associated with fewer emissions than three respiratory activities, and HFNO did not increase respiratory-generated aerosols.^20^ While comparisons were complicated by their different measures of peak and average counts, they too concluded cough was the most likely source of hazardous aerosols irrespective of therapies.^20^

These and other recent studies to quantify respiratory aerosols have collected particles at a short distance from the subject, using a small sampling inlet in free space with very low air sampling rates, typically of 1-5L/min.^19,20,25–27^ Accurately sampling exhaled aerosols is made challenging by airflows in excess of 600L/min, nebulous plumes travelling up to 60m/s and the addition of high volumes of ventilator gases.^28,29^ To avoid incomplete sampling by an unknown degree, we positioned the subjects’ heads within a large cone and sampled at 100L/min, attempting to capture as close to total emissions as possible. This novel approach could explain why our study demonstrates such markedly increased overall fold differences compared to prior studies of both activities and therapies.^19,20,26,27^

Our study is consistent with several others in demonstrating respiratory activity and exertion dramatically generate aerosols, which increase with speech loudness, greater breathing rate and volume and particularly during coughing.^24–26,30–32^ From the perspective of the physiological factors involved, the increases with activities are associated with rises in subglottic pressure, aerodynamic shear stresses and vocal fold and terminal airway open-closure frequency.^9,25,29,32,33^ In contrast, the pressure changes and flow velocities generated during respiratory therapies are far less.^28,34,35^ The slight increases with flow and pressure in emissions during breathing with HFNO may be due to turbulence within the upper airways, whereas NIPPV could generate increases through greater tidal volumes and subsequent airway open-closure, overriding our benchmark measure of ‘quiet breathing’. The increase in emissions with NIPPV-D may be explained by a greater degree of ventilator-subject desynchrony, which we speculate, may create pressure spikes within the facemask, causing leaks and aerosol generation at the mask-skin interface.^9,36,37^ All these increases are small compared to those with respiratory activities and are likely only detectable because of our sampling system and very low background counts. Our study suggests the physiological benefits of positive airway pressure, which splints open airways and reduces the pressure changes required to breath efficiently may reduce aerosols.^15,34,35^ The inclusion of exercise and forced expiratory manoeuvres, as proxies for symptomatic laboured breathing and atelectasis, suggests an aerosol-suppressing role for such therapies in patients with respiratory distress.

When a surgical mask was worn, the apparent filtration increased during higher velocity activities, however our video study suggests this was partly due to masks deflecting gas away from the collecting cone. This is consistent with other studies also showing sideways leakage with surgical masks.^38,39^ While this deflection may blunt the forward plume and remove large droplets, reducing direct person-to-person exposure, aerosols could still accumulate in poorly ventilated spaces.^15^

Our study’s strengths are that we were able to capture most of the total particles emitted over the relevant size range 0.5 to 25 µm during both respiratory activities and therapies, with negligible background contamination. By visualising the complex exhaled airflow patterns, we increase the confidence our method captured respiratory aerosols during activities and therapies. Our analysis gives context by demonstrating the proportion of larger, but far less numerous sized particles. The estimations of total volume serve as a unique unifying comparator between activities, therapies and subjects and enables future risk modelling during numerous scenarios.

There are several limitations to our study. We recruited ten healthy subjects who performed a single protocol run. There may have been some differences in how subjects performed activities, limiting the interpretation of the previously observed wide inter-subject variation.^25,30^ We estimated the average residence time for particles in the cone should have largely been sufficient for them to reach equilibrium diameter.^40^ However, given wide variation between activity airflows, volumes and the addition of humidified therapy gases, we are unsure exactly what proportion of variation in size distributions is physiological or methodological. Our post-hoc study shows cough and FEV are modestly incompletely sampled, highlighting the most aerosol generating activities are also the most challenging to comprehensively measure due to high airflow velocities. Our attempts to model acute respiratory physiology and symptoms with volitional activities are likely to differ from patients with COVID-19. However, Hamilton observed a comparable skew in particle size distribution in both COVID-19 patients and healthy controls suggesting potential similiarty.^20^ Crucially, all studies measuring aerosols are limited by not quantifying viable virus. Future work is needed to establish if physiological exertion and respiratory symptoms increase total viral and aerosol emissions in patients, as our study suggests.

Our data suggest the historic epidemiological association between the use of respiratory therapies and SARS-CoV-1 transmission should be reconsidered. The therapies were indicated for acute respiratory failure, and will have been associated with healthcare workers spending prolonged periods in close proximity to physiologically deteriorating patients with fast, deep breathing, cough and terminal airway closure.^13^ Our data suggest these activities may have resulted in large numbers of physiologically generated aerosols. Paradoxically, the introduction of respiratory therapies may modestly supress these. It seems likely, if aerosols were responsible for disease transmission, they were physiologically, not procedurally generated. This distinction is important as aerosol protective measures are currently prioritised based solely on procedure.^3,4^

Our data adds to a small but growing number of quantitative studies to challenge the rationale for describing certain respiratory therapies as AGPs.^17–20^ Patients acutely requiring HFNO or NIPPV are likely to present a high disease transmission risk, but we find no basis for withholding or delaying access to these therapies. We conclude instead, that respiratory activities are the primary modes of aerosol generation and represent a greater transmission risk than is widely recognised. Therefore, increased measures targeting physiologically generated aerosols could protect patients, healthcare workers and the general public from respiratory pathogens, including SARS-CoV-2.

## Data Availability

Share upon reasonable request.

## Acknowledgments

We thank the subject volunteers for their participation, the entire Intensive Care Department at the Prince of Wales Hospital for their support and patience. For invaluable assistance reviewing the manuscript we thank: Mark Jermy, Ph.D, Alistair McNarry, F.R.C.A., Karim Brohi, Ph.D, Justin Morgenstern, M.D, Donald Waters, M.B Ch.B., David Bihari, F.R.C.P., Moira Johns, M.R.C.P., Mo Theaghlach, Ph.D, David Carruthers, M.B., B.S., Pippa Dwan, M.B., B.S., Ruaridh McCusker, M.B Ch.B., Grace Boos, LLB., Rebecca Rowley, M.B., B.S., Anthony Thaventhiran, M.B., B.S.,

