## Supplementary Appendix for "The effect of respiratory activity, ventilatory therapy and facemasks on total aerosol emissions"

This appendix has been provided by the authors to give readers additional information about their work.

Table of Contents

*1. Supplemental Methods* ... 4
1.1 Detailed Description of Sampling Chamber. ... 4
1.2 Statistical Analysis Plan. ... 6
1.3 Post Hoc Visible Exhaled Aerosol Airflow Study ... 7
1.4 Post Hoc Comparison of Ventilator Pressures and Flows during the Delivery of Non-Invasive Positive Pressure Ventilation. ... 8
1.5 Ventilator Configuration ... 10
1.6 Additional Medical Equipment Information and Instruction for Use. ... 12
1.7 Model of Particle Size to Volume. ... 13
1.8 Relative Humidity, Temperature and Particle Equilibrium Diameter ... 14
*2. Supplemental Figures and Tables.* ... 16
Figure S1 Fold Changes in Particles with Different Aerosol Generating Procedures and Masks. ... 16
Figure S2 Total Volumes of Particles Generated by Six Different Activities by each Subject. ... 17
Figure S3 Visual Demonstration of Exhaled Particle Capture. ... 18
Video S1 Post-Hoc Visible Exhaled Aerosol Airflow Study Video. ... 20
Table S1 Design of Study. ... 21
Table S2 Particle Size to Volume Conversion Table. ... 22
Table S3 Subject Baseline Characteristic Data. ... 23
Table S4 Temperature and Relative Humidity. ... 24
Table S5 Physiological Data during Exercise whilst Receiving Non-Invasive Positive Pressure Ventilation. ... 25
Table S6 The Distribution of Particle Number and Total Sum of Particle Volumes. ... 26
Table S7 Fold Difference in Particle Counts when Compared to Breathing Alone across all Subjects and when the Highest is Removed. ... 28
Table S8 Recorded Ventilator Parameters for Varying Inspiratory Pressures in Two Subject Post-Hoc Study ... 29
References. ... 31

**Investigators:**

[1, 2] Nick M. Wilson, M.B., B.S.,
[3] Guy B. Marks Ph.D,
[1] Andrew Eckhardt M.B., B.S.,
[4] Alyssa Clarke M.Sc
[1] Francis Young, B.Med., M.D.,
[5, 6] Frances L. Garden Ph.D
[1] Warren Stewart B.Nurs
[7,8] Tim M. Cook, M.B., B.S.,
[9] Euan R. Tovey, Ph.D

**Institutions:**

[1]Department of Intensive Care Medicine, Prince of Wales Hospital, Sydney, Australia
[2]Department of Anaesthesia, Royal Infirmary of Edinburgh, NHS Lothian, Scotland, UK
[3]University of New South Wales, Sydney, Australia
[4]Department of Intensive Care, Royal Prince Alfred Hospital, Camperdown, NSW, Australia
[5]South Western Sydney Clinical School, University of New South Wales, Sydney, Australia
[6]Ingham Institute of Applied Medical Research, Sydney, Australia.
[7]Department of Anaesthesia and Intensive Care Medicine, Royal United Hospital NHS Trust, Bath, UK
[8]Bristol Medical School, University of Bristol, UK
[9]Woolcock Institute of Medical Research, University of Sydney, Glebe, NSW, Australia

**Acknowledgments:**

We thank the subject volunteers for their participation, the entire Intensive Care Department at the Prince of Wales Hospital for their support and patience. For invaluable assistance reviewing the manuscript we thank: Mark Jermy, Ph.D, Alistair McNarry, F.R.C.Anae., Karim Brohi, Ph.D, Justin Morgenstern, M.D, Donald Waters, M.B Ch.B., David Bihari, F.R.C.P., Moira Johns, M.R.C.P., Mo Theaghlach, Ph.D, David Carruthers, M.B., B.S., Pippa Dwan, M.B., B.S., Ruaridh McCusker, M.B Ch.B., Grace Boos, LLB., Rebecca Rowley, M.B., B.S., Anthony Thaventhiran, M.B., B.S.,

### 1. Supplemental Methods

#### 1.1 Further Description of Sampling Chamber and Protocol

The chamber (see Figure 1) was 130 x 60 x 60 cms overall and partitioned into a rear smaller pressurised chamber containing a domestic air filter (Oblozone Air Purifier, Medical Grade, H13 True HEPA, Australia) and a forward chamber which contained the torso of the subject and the sampling cone. Because the air filter alone would not provide sufficient airflow, a continuous positive airway pressure (CPAP) device (Sullivan Autoset, ResMed, San Diego, United States) was used to pressurise the rear chamber via a respiratory filter (Suregard, RJVKB, Bird Healthcare, Melbourne, Australia). Filtered air flowed from the rear chamber into the subject chamber through a partition wall consisting of two layers of the aerosol filtering material, (electret, ETR 115, Vilene, Japan) which also functioned as a flow distributor. Airflow in the 100 mm diameter tube at the tapered distal end of the sampling chamber was 0.6m/sec, equivalent to 72 cm/min flow across the chamber; this reduced to ~0.4 m/sec when the optical particle counter (OPC) was sampling at 100L/min, as this also removed air from the chamber. Airflows were validated by video recordings of aerosols from an e-cigarette.

The occupied portion of the chamber has clear walls and the base contained a waterproof nylon skirt with a drawstring, allowing the torso of the seated subject and tubing associated with the ventilation methods to be sealed into the chamber. Samples were collected using a cone of total length 80 cm, maximum width 35 cm (volume 27L) with a 30 cm portion of the side wall removed so the subject's head fitted inside, such that the head was surrounded at the top and sides. The resultant volume of cone in front of the head was approximately 13 L. The cone was made of electrically conductive aluminium-coated cardboard and connected by 33cm of 0.95 cm ID siliconized tubing to the laser OPC located outside the chamber.

During the study, subject heart rate, pulse oximetry and ventilator variables were monitored continuously. The relative humidity and temperature in the box and the cone were monitored during some runs. Measurements within the chamber used the following instrumentation: within the chamber, temperature and relative humidity with Hygrometer Thermometer (KT-905, DealMux, China), airflow in the exhaust tube (Hot wire Anemometer, WD9830, HTi, Dongguan Xintai Instruments, China), and temperature and relative humidity within the sampling cone (Temperature and Humidity Meter HT-350, HTi, Dongguan Xintai Instruments, Dongguan City, China).

During the development of the chamber, it became apparent that non-respiratory aerosols were a potential source of error. Sources of these included: subject's hair, skin, clothing, any ambient environmental air (and medical gas supplies. To mitigate against these contaminants, we designed and developed the chamber described above so the overall pressure within the chamber was higher than the environmental pressure to prevent ambient air entrainment. This was enabled by the overall airflow through the air filter to exceed the 100L/min sampling from the OPC. When a subject first settled into the chamber the counts were extremely high, reflecting external air that was brought into the chamber, but also within the subject's lungs.Sampling did not commence until this washout had concluded, normally taking around 2 minutes.

Equipment was another potential source of contaminant error. We placed the surgical masks, HFNO and NIPPV within the sample cone, both physically within the cone and a separate experiment on a polystyrene manikin head and performed experimental sample runs. Aerosols were initially generated during HFNO, which we thought might be related to the humidity or moisture, but the source was established to be the supplied gas as medical air contains environmental particles. When an additional filter was placed on the HFNO circuit this contamination was removed and HFNO did not produce intrinsic aerosols. We were again able to demonstrate during NIPPV there was no intrinsic aerosol generation. Although we were unable to test the pressure delivery mode as this requires an initiated breath, but on continuous positive airway pressure mode no aerosol contamination was noted.

To minimise subject contamination and mask leakage, we asked for males to shave on the day of the protocol. Subjects were given a newly opened polypropene hooded jacket to wear, they were asked to keep still during the protocol and look directly ahead, to minimise clothing or hair disturbance. We were concerned during exercise the motion of the legs, or general movement of the torso could give a false aerosol reading, so we recreated this level of movement, but wearing an N95 respirator and noted no contamination signal. It is likely any contaminant aerosols entering the box from around the skirt are transported beneath the cone in line with the flow direction and therefore not sampled. This is supported by our vape smoke videos of airflow within the chamber and by the increase in contamination when the air pump flow rate is reduced. On the established experimental settings, the flow rate provided surrounded the cone with clean laminar airflow which prevented contamination from entering the sampling cone. To give added assurance that we were truly sampling respiratory aerosols one of the subjects performed the entire respiratory therapy protocol whilst apnoeic. They had prior breath hold experience and held their breath for one-minute intervals, during sampling, whilst receiving respiratory therapies. Again, no signal of aerosol generation was observed during these un-published development studies.

. To increase our confidence in the protocol one of the subjects performed the protocol but whilst doing replicates of 3s, all 1-minute samples. This unpublished data showed a reassuring consistency between samples.

#### 1.2 Statistical Analysis Plan.

The primary outcome was particle count, classified into six size bins with cut-points at 0.5µm, 0.7 µm, 1.0 µm, 3.0 µm, 5.0 µm and 10 µm. As the counts were positively skewed and included some zero values, the counts were transformed to an approximately normal distribution by the formula: log-count = $\log_{10}(count + 0.3)$ .

The primary objectives were to estimate the effect on particle count of:

1) Various activities, compared to quiet breathing, on particle count.
2) Various respiratory therapies during varying modes of use and surgical facemasks.

Secondary objectives were:

3) To test whether there was any interaction between activities, respiratory therapies and surgical facemasks in their effect on particle count.
4) To estimate whether the effect of activities, respiratory therapies and surgical facemasks had divergent effects on particles of various sizes.

The analysis was conducted using mixed effects regression models (in Proc Mixed, SAS version 9.4). In all models the dependent variable was log-count. For models investigating the effect of activities the main fixed effect was activity. For models investigating the effect of facemasks on activities the main fixed effects were activity with and without facemasks. For models investigating the effect flow rate in HFNCT (High-Flow

Nasal Cannula Therapy), the main effects were activity and flow rate. For models investigating the effect of type of NIPPV (non-invasive positive pressure ventilation) and IPAP (inspiratory positive airway pressure) pressure, these were included as main effects with activity and pressure. Table S1 details the various combinations of activity, therapy and facemasks used in the models. Reference values were quiet breathing (for activity), no respiratory therapy (for respiratory therapy) and no surgical mask (for surgical mask).

The complex repeated measures design required a complex analysis. We treated the multiple measures of log-count (corresponding to the six size bins) as repeated measures using a random intercept. Hence, we included two random "subject" statements: one to account for repeated measures within participants and the other to account for repeated measures of particle count. In both random statements we included random intercept and covariate terms (the latter allowing for random slope on the factor of interest). We assumed a variance components covariance structure. Degrees of freedom for the estimation of standard error of the fixed effects was estimated using the Kenward Roger method.[1] Estimation was by the Restricted Maximum Likelihood (REML) method. The comparisons between the activities and surgical facemasks and/or respiratory therapies respiratory activities are reported as a fold difference (with 95% confidence intervals) from the geometric mean of the activity, respiratory device to their reference.

#### 1.3 Post Hoc Visible Exhaled Aerosol Airflow Study

**Method:**

A transparent cone was constructed from sheets of clear acrylic film of the same dimensions as the aluminium-cardboard cone used in the main study. Air was drawn from the cone, to simulate the OPC sampler, by connecting it to the air inlet of a CPAP device (Sullivan C6, ResMed, San Diego, United States) and manually setting the flow at 100L/min (Flowmeter, model 4000, TSI Inc, Minnesota, United States). The cone was illuminated by three 100W halogen lamps, each about 90cm from the cone, two above and one from being the subject's head, with appropriate screens to confine the illumination to the area of aerosol dispersal. To maximise contrast of the aerosols, black cardboard sheets were used in the background, and the subject was dressed in a dark hooded sweatshirt.

Electronic nicotine delivery systems, commonly referred to as electronic cigarettes generate, propylene glycol and glycerin aerosols. The size of the aerosols has been measured elsewhere to have peak in size distribution around 250-450 nm depending on the actual device, flow rate and other factors.[2] Thus, its flow reflects aerosols not droplets. We selected them for use due to their safety profile, easy visualisation and aerosol size. The e-cigarette device used was a Vinci Air (VAPOO, Guangdong, China) using the 1-ohm element on the 15W setting. Nicotine-free 'juice' was composed of propylene glycol, glycerine, and artificial flavour (Nimbus Vapour, Marrickville, Australia). A Fujifilm digital camera X-T2 Body firmware version 4.20 with XF 18-55mm f/2.8-4 Lens firmware version 3.22 (Fujifilm, Tokyo, Japan) was used in movie mode to record the experiments, with the video edited in iMovie version 10.2.2 (Apple, Cupertino, California, United States).

Two subjects from the main study were recruited; a male and female aged 26. The results are shown in Video S1 - Post-Hoc Visible Exhaled Aerosol Airflow Study Video. Each participant performed the activities breath, talk, shout, FEV and cough three times, attempting to replicate the patterns used in the study. However, owing to the adverse effects of the flavoured aerosols on speech and respiration, this was not always achieved. The respiratory activities were repeated while wearing the same surgical mask from the mains study, which was moved aside to enable inhalation of vapour and then put back on for the exhalation. It difficult to perform the experiments during the respiratory therapies HFNO and NIPPV due to technical challenges and risk of equipment damage. Therefore, a limited number of recordings were made to characterise the exhaled airflow patterns, primarily to ensure that exhaled gas was sampled efficiently. These were performed with the same masks, ventilator settings as the main study. HFNO was performed at 60L/min, NIPPV-S and D were both performed at IPAP/EPAP 25/10 $cmH_20$. Exercise was excluded to prevent subject discomfort. No attempt was made to quantify the vape smoke and these videos are indicative and instructive rather than quantitative. Video is available in the Figure section.

#### 1.4 Post Hoc Comparison of Ventilator Pressures and Flows during the Delivery of Non-Invasive Positive Pressure Ventilation.

**Method:**

To compare performance subject interactions and characterise total flows and leaks a post hoc study was performed. Two subjects from the main study were recruited; a male and female both aged 26. The ventilator protocol from the main study was used, whereby subjects received incremental increases in pressure of 5 $cmh_20$ sequentially until they received 25/10 IPAP/EPAP. Exercise was performed on a pressure setting of 20/10 cm h20. The study was not performed in the aerosol measuring chamber and aerosols were not sampled. Each ventilatory pressure change was maintained for 1 minute, during which a video recording was taken of the ventilator screen. During this minute of recording, the 3 most similar waveforms were chosen to gather data for each pressure interval. Recorded parameters were displayed on the ventilator screens during the minute of measurement. Waveforms were analysed over the minute of recording by the investigators to estimate an asynchrony index (ASI). Gross asynchronous events/breaths (double triggering, ineffective triggering, autotriggering) were divided by the displayed total respiratory rate (per minute) and multiplied by 100 to give a percentage. The results were analysed using the paired t-test. We deemed high levels of asynchrony if ASI>10%. [3,4]

**Results:**

Peak inspiratory and expiratory flows were largely similar between the two ventilators (see Table S7.). The maximal leak and minute volumes during exercise did not exceed the OPC sample rate of 100L/min. The NIVPPV-D C6 Hamilton was associated with a higher mean respiratory rate (15.4 versus 13.7 per minute, p=0.03), tidal volumes (2.2 versus 1.75 L, p=<0.001) equating to a 23% greater minute ventilation. The V60 single circuit NIPPV-S had a large expected leak, which increased with pressure and exercise, to reach a maximum of 30 litres per minute during 20/10 exercise. The closed C6 dual circuit had a smaller leak, peaking at 3 litres per minute during 20/10 quiet breathing, but was consistent and present at lower pressures. Given the C6 circuit was enclosed, this leak was presumably occurring at the patient-mask interface and via the anti-asphyxia valve. Leaks tend to exacerbate asynchrony particularly in an enclosed dual circuit system.[3,4] The ventilators were not adjusted in the case of asynchrony. Leaks were reported subjectively as perceived high velocity airflows from the mask. An increased number of asynchronous events were experienced on the C6 ventilator (ASI 29.8) compared to the V60 (ASI 1) which was also appreciated subjectively by the participants. Most asynchronous events were double triggering. For recorded results refer to Table S7.

#### 1.5 Ventilator Configuration

Both ventilators were configured as per current clinical practice at the institution where the study was undertaken.

**The C6 Ventilator** (Hamilton, Bonaduz, Switzerland) was used with a 950 humidification unit (Fisher and Paykel, East Tamaki, Aotearoa) and a Expiratory Heater Filter (800-VH330, VADI, Taoyuan City, Taiwan). The Fisher and Paykel 950 Adult Ventilator Dual Heated Circuit Kit (950A81) was attached to the ventilator and humidification unit with the included RT019 filter on the C6 inspiratory port. A further Fisher and Paykel RT019 filter was placed on the end of the flow sensor at the patient end. A Fisher and Paykel Nivaro RT045 type mask was fitted by an intensive care doctor as per manufactures instructions and connected into the RT019 filter to complete the circuit. The subjects were in an enclosed chamber without ready hand access to their face, to prevent risk in the event of gas supply failure during exercise the mask had an anti-asphyxia valve. A 2000ml 'Uromatic' 0.9% Sodium Chloride fluid bag (Baxter, Illinois, United States) was used for the humidification unit. Settings used on the 950 humidification unit were 'mask' function at 33˚C.

The NIV (non-invasive ventilation) mode is an implementation of non-invasive positive pressure ventilation (NIPPV). The pressure support (Psupport) setting defines the applied pressure during inspiration. NIV mode was programmed for flow cycled breaths. Every patient inspiratory effort trigger results in a flow-cycled, pressure-supported breath. Expiratory trigger sensitivity (ETS) defines the inspiratory timing of the breaths. Intellisync was in use as an inspiratory and expiratory flow waveform analysis software designed to minimise patient-ventilator asynchrony. To synchronize, IntelliTrig compensates for leaks and resistance between the ventilator and the patient, and with each breath, it measures the leakage at the patient interface (mask). With this information, IntelliTrig adjusts the trigger mechanism, reducing the influence of leakage and the changing breath pattern on the operator-set trigger sensitivity. Significant leakage can be present which can serve to reduce the actual applied PEEP/CPAP and give rise to auto-triggering. An anti-asphyxia mask is not classically used with the NIPPV-D sealed circuit as it may exacerbate ventilator desynchrony through leakage that can be interpreted as a breath. If the ventilator does not detect an expiratory trigger (for example, due to a leak), inspiratory time is limited by TI max.[5,6] After selecting subject age, sex, height, the required PEEP and PS, the settings used on the C6 ventilator were as follows:

- Mode: NIV

- P-ramp: 50 milliseconds
- TI max: 2.0 seconds
- ETS: 25%
- Oxygen: 25% Fi02
- Cycle: Intellisync+
- Trigger: IntelliTrig flow waveform trigger

**The V60 Ventilator** (Phillips, Eindhoven, The Netherlands) was used with a Fisher and Paykel 950 Humidification unit and a Fisher and Paykel Adult 950 Adult Bi-Level/CPAP Heated Circuit Kit (950A61). On the ventilator outlet a bacterial/viral filter (303EU, Vyaire, Mettawa, United States) was used. The circuit was completed with a RT019 filter on the end of the long circuit limb, a RT017 exhalation port attached to the filter and the fitted RT045 Nivaro mask attached to the exhalation port. Humidification settings and fluids were the same as the C6 configuration.

Mode used was S/T (Spontaneous/Timed). The S/T mode delivers breaths at the user-set rate. It delivers pressure-controlled, time-cycled mandatory and pressure supported spontaneous breaths at the IPAP pressure level. If the patient fails to trigger a breath within the interval determined by the Rate setting, the ventilator triggers a mandatory breath with the set I-Time.[7,8] The patient triggers and cycles based on the ventilator's Auto-Trak Sensitivity software. After selecting the required IPAP and EPAP (during this experiment, the 'CPAP' mode was not used, instead, both the IPAP and EPAP values were set to the same number in S/T mode) the following settings were used:

- Rate: 4 BPM
- I-time: 1.2 seconds
- Rise: 1
- Oxygen: 25% Fi02
- Ramp: Off
- Cycle: AutoTrak sensitivity software
- Trigger: AutoTrak - flow waveform triggered

For further technical details refer to the V60 Ventilator User Manual.[7]

#### 1.6 Additional Medical Equipment Information and Instruction for Use.

**Non-Invasive Ventilation Facemask and High-Flow Nasal Cannula Fitting Instruction** [9,10]

Medical device used for Non-Invasive Ventilation: Fisher and Paykel Nivaro RT045 Non-vented Hospital Full Face Mask Anti-Asphyxiation Valve Version. Non-vented NIV masks with an anti-asphyxiation valve do not have integrated leak ports and are designed for use with single limb breathing systems which have a CO2 leak integrated into the circuit port. The CO2 expiratory leak port in the breathing system removes CO2 during the exhalation phase. These masks have a safety feature, with an anti-asphyxiation valve that opens in the case of an unexpected low supply pressure and ventilator failure, or breathing system disconnection, allowing the subject to breathe spontaneously within the measuring chamber.

Medical device used for High-Flow Nasal Cannula: Fisher and Paykel Optiflow Plus (size medium). Product code OPT944

Fitting instructions: As per the measurement card and instructions provided by the manufacture. [6-7]

**Surgical Facemask Fitting Protocol, Standard and Equipment Used**

Medical device used: MedCon Surgical Face Mask (Shepparton, Australia). Code 170515. ARTG no 109872. Built to AS 4381:2015. Bacterial filtration efficiency ≥98%. Differential pressure <5.0 mmH2O/$cm^2$. Resistance to penetration by synthetic blood (minimum pressure for pass result) 160mmHg. Note particulate filtration efficiency is not required for this standard.

Fitting instructions: Surgical masks were fitted as per the Royal Australian College of General Practitioners (RACGP) guideline.[11]

#### 1.7 Model of Particle Size to Volume.

The TSI AeroTrak® Portable Particle Counter Model 9500 (TSI instruments, Minnesota, USA) provides data in six size bins, 0.5–0.7, 0.7–1, 1–3, 3-5, 5-10, and 10-25 µm to reflect skewed particle distribution.[12]

In the literature, the shape of the distributions for particle size and numbers differs, and depends on the activity, the precise methods for determining size and allowances for reaching equilibrium diameter.[13,14]

The simplest option was to presume a flat distribution of sizes within each bin.

In our study we presumed, based on sampling rate and volume of the cone, and adequate mixing with ambient air with the time between emission and measurement being >2 seconds, that particles should have reached equilibrium diameter.

The average volume of a particle within each bin was calculated by averaging the volumes of 5 equally spaced sub-bins. The volume of a sub-bin was defined as 4/3*pi*radius^3, where the radius of the sub-bin was the average of the upper and lower radius of the sub-bin. Refer to Table S2 for particle size to volume conversion.

#### 1.8 Relative Humidity, Temperature and Particle Equilibrium Diameter

In these experiments, particles were expelled from either the mouth or ventilation devices within a plume of saturated warm air; this plume had a direction and velocity depending on the experiment. As this plume mixes with the surrounding cooler and drier ambient air, the particles of respiratory fluid will rapidly dehydrate until they reach an equilibrium diameter. The final diameter is mainly determined by the resultant temperature and humidity of air mixture plus the proportions of the non-aqueous components of the fluid droplet. The time required to reach the equilibration diameter is a function of the particle's original size, the temperature, humidity, and turbulence of the ambient air.

Our understanding of equilibrium diameter is mainly based on theoretical models that make several assumptions, which have been supported by observations. Higher ambient RH slows the dehydration process. The well-cited model by Nicas (2005) proposed the equilibrium diameter would range from 0.47 to 0.61 of the original, for percent relative humidity (RH) in the range of 30% to 60%, with an estimated time to reach final diameter of 10 µm of between 0.17 and 0.4 seconds.[15] Smaller particles equilibrate much more rapidly. More recent models give slightly greater decreases in diameter. For example, Stadnytskyi (2020), citing pre 1946 references, and without reference to humidity, gives the final diameter of salivary droplets to be 'about 20 to 34% of the original size', whereas Netz (2020) gives the typical shrinkage to be 0.33 of the original diameter at 50% RH;  for a particle of initial diameter of 10 µm, this requires a time of 0.5 seconds, while a 20 µm particle shrinks to 7 µm in less than 2 seconds at 50 % RH.[16,17] These values are not inconsistent, given the varying assumptions on humidity and non-volatile content.

We recorded data for Data for % RH and temperatures in the chamber for seven-ten subjects across the protocol and shown in table S4. The differences in ambient RH were small and were determined by environmental variation, whereas indoor temperatures were relatively stable. There was little evident increase in RH in the chamber itself during respiratory therapies, probably due to most expelled air going down the cone and the slight increase in temperature was probably associated with human occupancy of the chamber and heat from the air filter pump.

As measuring the humidity and temperature within the cone could alter the deposition of particles and airflow within the cone we only performed ad-hoc simultaneous measurements of temperature and humidity within both the chamber and within the cone, as described in S1.1; see detailed description of the sampling chamber. We measured temperature and relative humidity within the cone with two subjects during the six respiratory activities, and one run of HFNCT and one run of NIPPV-S. The temperature within the cone changed by 0.5-3$^{o}$C with respiratory activities and by 1-3 $^{o}$C during NIPPV-S and HFNCT. The greatest increase of RH in the cone during activities was with exercise and forced expiratory volume manoeuvres, with increases from 29-30% to 60%, with NIPPB-S the maximum %RH was from 29.6% to 59.5% (average 52.8%), with HFNCT the maximum % RH increase was from 35% to 74.7 % (average 69%).

In our study the volume of the cone in front of the subject was at least 13 L, and the OPC flow rate 100L/minute. As shown in the accompanying videos, different patterns of exhalation into the cone result in different flows and turbulent mixing patterns; a small proportion of the exhaled aerosol probably could reach the inlet tube in around one second while the entire sample would require >10 seconds. The transit time in the inlet tube itself is ~0.2 seconds. Thus, almost all the aerosol, particularly particles <10 µm, reach equilibrium diameter long before they reach the OPC, presuming the mixed % RH is within the ranges commonly modelled.

Models would indicate that particles in our study would have sufficient time to reach their equilibrium diameters prior to entering the OPC. Differences created by outdoor conditions on the same day, would apply to all samples collected. Based on the model of Nicas, which hypothesises small increases in equilibrium diameter as humidity rises from 40% to 60%, it seems likely the relatively higher humidity created by some activities with high minute volumes, or therapies contributing warm moist air, might result in small increases in diameter. Detailed continual monitoring of the complex changes in humidity and airflows within the cone accurate to fractions of a second was beyond the capabilities of our instrumentation and an acknowledged limitation of our and other similar studies collecting exhaled particles.

#### 2. Supplemental Figures and Tables.

#### Figure S1 Fold Changes in Particles with Different Aerosol Generating Procedures and Masks.

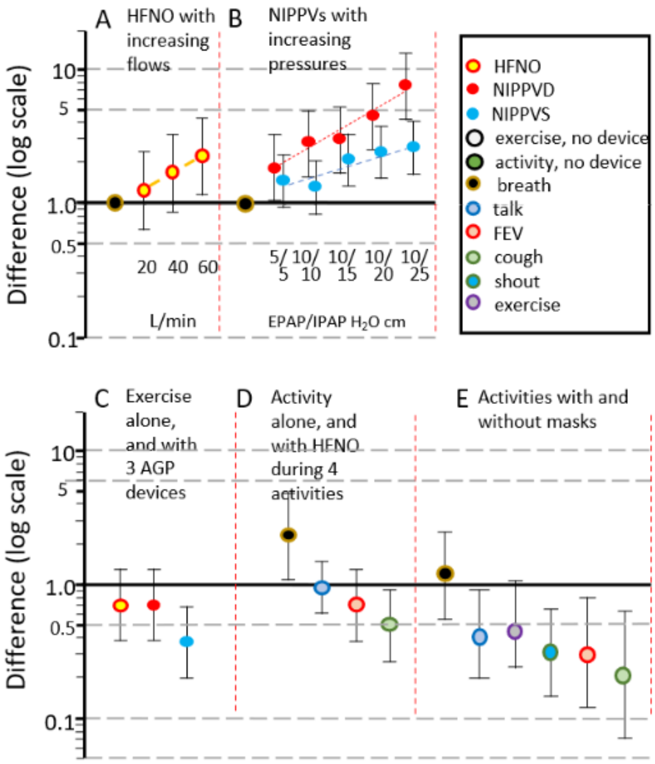

Figure S1: Shows fold changes in particles and 95% confidence intervals plotted on a log scale with different aerosol generating procedures (AGP) and masks. Fig 3A shows fold increase in particles, relative to quiet breathing, for high flow nasal canula (HFNO) at the flows shown in L/min. Fig 3B shows the fold change in particles relative to quiet breathing for both non-invasive positive pressure ventilation, single and dual circuits, NIVVP-S and NIVVP-D respectively at the EPAP/IPAP pressures shown, in cm $H_2O$. Fig 3C shows the relative fold change for the three different AGPs when used during exercise at their highest settings used, compared to exercise alone. Fig 3D shows the fold change for HFNO during four different activities relative to the same activity with no HFNO. Fig 3E shows the change in particle counts of six activities with masks, relative to the particle counts of the same activity without masks.

### Figure S2 Total Volumes of Particles Generated by Six Different Activities by each Subject.

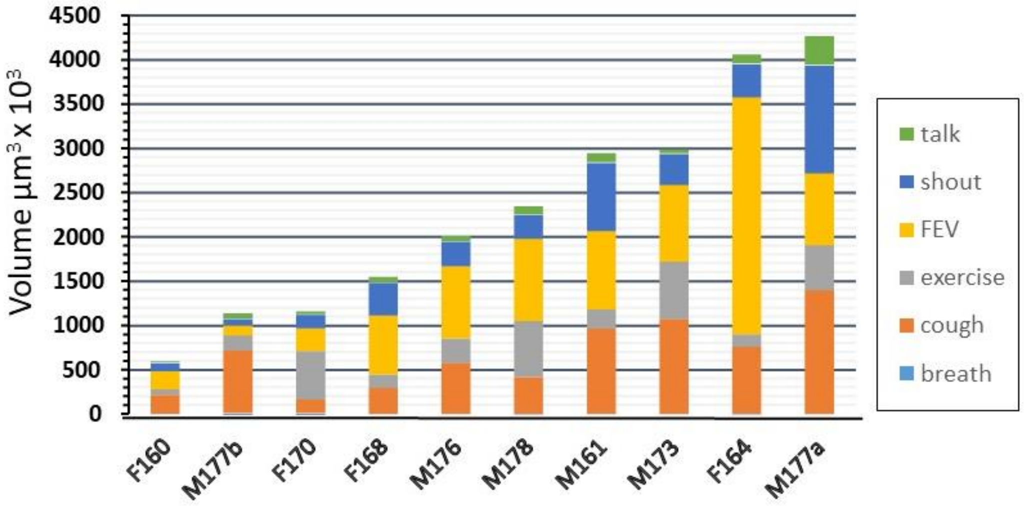

Figure S2. Total volumes of particles generated by six different activities by each subject. Subjects described by gender (M/F) and height (cm), two subjects were male and 177 cms. Volumes estimated using a simple model applied to the six size-bins of particle counts 0.5 to 25 µm. Quiet breathing contributes <0.2% of total volume on average and is barely discernible.

### Figure S3 Visible exhaled aerosols within sampling apparatus during respiratory activities and therapies

**Figure S3 1** – Photographs taken of a subject within the aerosol measuring chamber during the sampling protocol. Note, backwall made of electret material, Perspex walls, experimental sampling cone composed of electrically conductive aluminium coated cardboard, polypropene hooded jacket worn by subject. **Note – all photographs have needed to be removed due to Medrix policy. Please contact the author if you would like access to these images.**

**S3.2 -** Subject observed during talking, exhaled gases observed to be almost entirely sampled. Subject anonymity is maintained as per Medrix policy (1:25:08*)

**S3.3 -** High efficiency of particle capture during breathing receiving HFNO at 60L/min flow. Despite high exhaled gas dispersal due to HFNO flow, the large cone and high sample rate sampling maintains high sampling efficiency (2:55:05*).

**S3.4 -** High efficiency of particle capture during closed limb NIPPV-D IPAP/EPAP pressure 25/10 cmH20. Note leakage via the facemask anti-asphyxia valve. (3:08:17*)

**S3.5 -** High efficiency of particle capture during open limb NIPPV-S breathing IPAP/EPAP pressure 25/10 cmH20. Two exhalation plumes visible, the first via anti-asphyxia valve near to mask, the second via the open expiratory port towards bottom of picture (3:41:08*)

**S3.6 A-D** - Series of 4 images during the early phase of a cough recorded over 9 milliseconds. High velocity nebulous cough plume visible which undergoes rapid turbulent mixing within the cone (1:45:05; 1:45:08; 1:45:11; 1:45:16*).

**S3.7** – Image from 4 seconds following a cough. High velocity has gas is observed to be deflected backwards and an unquantified proportion is ejected from the rear of the cone. (1:49:06*)

**S3.8 -** As observed during cough, the velocity and volume of cough and FEV exceed the volume and rate of sampling leading to loss of exhaled particles from the rear of cone during FEV. This suggests under-sampling during both cough and FEV despite the high sampling rate and large cone (1:38:23*)

**S3.9 -** Talking demonstrated deflection around the mask, which is subsequently captured on a an upward thermal plume of body heat and carried upwards and out of the cone (2:19:27*).

**S3.10 –** Considerable quantities of high velocity exhaled gas is observed deflecting backwards following a cough (2:42:10*)

*Images obtained from Video S1 Visible Exhaled Aerosol Airflow study. Provided is the timing of the photo displayed in the video.

Video S1 Post-Hoc Visible Exhaled Aerosol Airflow Study Video.

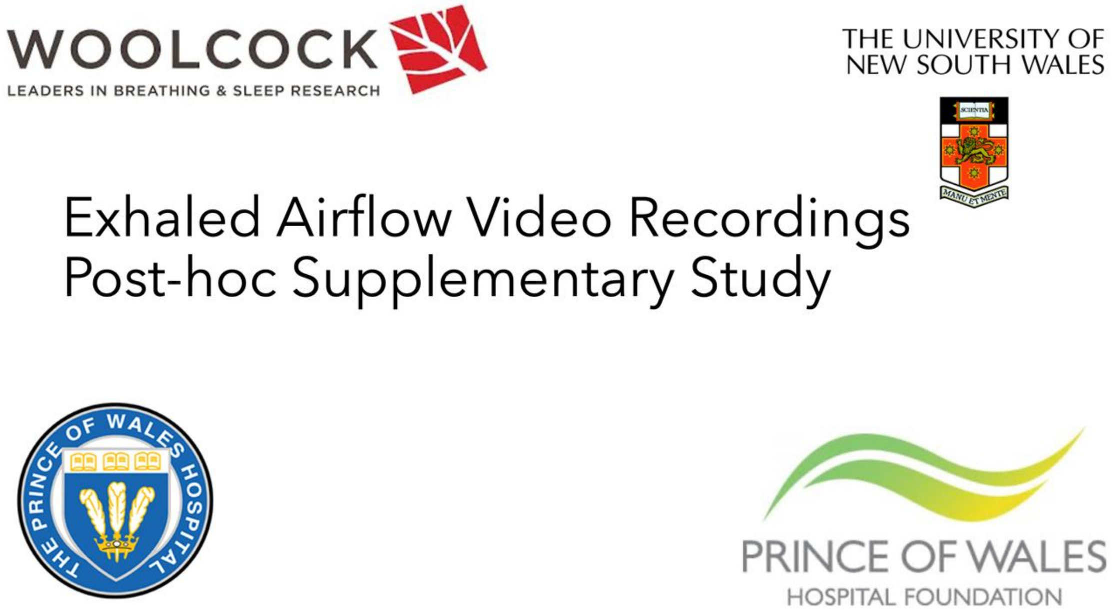

*Click on above icon to be redirected to video recording. Not available in Preprint.

### Table S1 Design of Study.

| | | Quiet breathing | Exercise | Cough | Forced Expiration | Talk | Shout |
|---|---|---|---|---|---|---|---|
| **No device** | | ✓ | ✓ | ✓ | ✓ | ✓ | ✓ |
| **Surgical mask** | | ✓ | ✓ | ✓ | ✓ | ✓ | ✓ |
| **HFNO*** | | ✓ 20, 40, 60 | ✓ 60 | ✓ 60 | ✓ 60 | ✓ 60 | ✗ |
| **NIPPV†** | **Single** | ✓ 5/5 to 25/10 | ✓ 20/10 | ✗ | ✗ | ✗ | ✗ |
| | **Dual** | | | ✗ | ✗ | ✗ | ✗ |

Table S1: Activities are shown in columns and devices in rows. Symbol ✓ indicates performed, ✗ indicates not performed. The study was a multi-way crossover design.

* Numbers are flow rates (L/min)

† Combinations of EPAP / IPAP pressures used with breathing were 5/5, 10/10, 15/10, 20/10, 25/10, in cm $H_2O$

Abbreviations: HFNO; high flow nasal oxygen. NIPPV; Non invasive positive pressure ventilation.

### Table S2 Particle Size to Volume Conversion Table.

| Lower diam | Upper diam | Av µm$^3$ |
|---|---|---|
| 0.50 | 0.70 | 0.116 |
| 0.70 | 1.00 | 0.331 |
| 1.00 | 3.00 | 5.192 |
| 3.00 | 5.00 | 35.505 |
| 5.00 | 10.00 | 244.348 |
| 10.0 | 25.00 | 3,299.515 |

Table S2 was used to convert particle number to particle volume. TSI AeroTrak® Portable Particle Counter Model 9500 provides data in six size bins, 0.5–0.7, 0.7–1, 1–3, 3-5, 5-10, and 10-25 µm. The simplest option was to presume a flat distribution of sizes within each bin. The average volume of a particle within each bin was calculated by averaging the volumes of 5 equally spaced sub-bins. The volume of a sub-bin was defined as 4/3*pi*radius^3, where the radius of the sub-bin was the average of the upper and lower radius of the sub-bin.

### Table S3 Subject Baseline Characteristic Data.

| Characteristic | Mean (SD) |
| --- | --- |
| Age (years) | 29 (2.83) |
| Height (cm) | 170.4 (6.88) |
| Weight (kg) | 71.4 (8.88) |
| BMI (Kg/$m^{2)}$ | 24.51 (1.75) |
| Resting HR (BPM*) | 63.7 (8.2) |
| Resting RR (PBM) | 11.77 (2.28) |
| FEV1 (litres) | 3.99 (0.41) |
| FVC (litres) | 4.84 (0.48) |
| FEV1/FVC (litres) | 82.85 (3.53) |

Table S3: Subjects included in the study comprised of 4 Females and 6 Males. SD denotes, standard deviation; cm, centimetres; kg, kilograms; kg/$m^{2,}$ kilograms divided metres squared, BPM*, breaths per minute; BPM, beats per minute; FEV1, forced expiratory volume in 1 second; FVC, forced vital capacity; FEV1/FVC, forced expiratory volume in 1 second divided by forced vital capacity.

### Table S4 Temperature and Relative Humidity.

| Activity | Temperature (°C) | | Relative Humidity (%) | |
|---|---|---|---|---|
| | **Mean (SD)** | **Range** | **Mean (SD)** | **Range** |
| Ambient | 23.5 (1.6) | 21.0 -26.0 | 37.5 (5.9) | 26.0-46.0 |
| No device | 26.8 (1.7) | 22.3-28.2 | 37.1 (8.4) | 28.0-52.0 |
| HFNO | 27.7 (1.3) | 26.0 -29.5 | 36.0 (5.1) | 30.0-46.0 |
| NIPPV-D | 28.1 (1.7) | 24.3-29.6 | 34.2 (4.6) | 28.0-42.0 |
| NIPPV-S | 27.8 (2.3) | 22.9-29.5 | 35.1 (3.9) | 29.0-40.0 |

Table S4: Recordings of temperature and relative humidity from within the chamber during the title experiment. Data sets from 7-10 subjects. Ambient conditions were recorded without a subject within the chamber. No device includes surgical masks and no mask. SD denotes, standard deviation; HFNO is high flow nasal oxygen, NIPPV is non-invasive positive pressure ventilation, either single circuit (S) or dual circuit (D).

#### Table S5 Physiological Data during Exercise whilst Receiving Non-Invasive Positive Pressure Ventilation.

| Physiological Parameter | C6 Hamilton Ventilator Mean (SD) | V60 Phillips Ventilator Mean (SD) |
|---|---|---|
| Minute Volume (L/min) | 29.50 (5.51) | 33.21 (10.29) |
| Tidal Volume (Litres) | 1.34 (0.40) | 1.49 (0.40) |
| Respiratory Rate (BPM*) | 22.14 (6.07) | 22.13 (5.51) |
| Heart Rate (BPM) | 119.00 (14.43) | 117.50 (15.64) |

Table S5: Recordings made during the title experiment using the displayed values on the ventilator at the end of the minute sample time. Performed with 20/10 cm $H_20$ Inspiratory Positive Airway Pressure/ Expiratory Positive Airway Pressure. Minute ventilation, tidal volumes and respiratory rates were recorded using the ventilators. A separate pulse oximeter was used to take heart rate readings. All values were recorded at the end of the exertion effort. Subjects were sporadically omitted from data inclusion for some of the pressure levels. Therefore, this is incomplete data and illustrative of the conditions during the study, but not definitive.; BPM, Breaths per minute; BPM, Beats per minute; L/min, Litres per minute.

* Respiratory rate measured in breaths per minute.

#### Table S6 The Distribution of Particle Number and Total Sum of Particle Volumes.

| **Activity or Therapy** | **Sum of all particle counts** | **Percent of particles <5µm** | **Volume of all particles (µm$^3$)** | **Percent of volume in particles <5µm** |
|---|---|---|---|---|
| Quiet breathing | 140 | 97.46 | 3,737 | 15.20 |
| Talk | 5,317 | 98.69 | 88,039 | 22.13 |
| Exercise | 5,591 | 94.76 | 336,888 | 5.94 |
| Shout | 29,970 | 98.86 | 393,108 | 21.69 |
| FEV | 68,447 | 98.15 | 826,651 | 14.53 |
| Cough | 140,370 | 99.50 | 653,972 | 34.89 |
| Mean* | 49939 | 97.9 | 459,732 | 19.8 |
| HFNO BR 20 | 122 | 94.51 | 9,749 | 8.83 |
| HFNO BR 40 | 259 | 96.22 | 12,089 | 7.13 |
| HFNO BR 60 | 609 | 97.15 | 8,187 | 22.41 |
| Mean | 330 | 95.96 | 10,008 | 12.79 |
| NIPPV-S BR 5/5 | 187 | 96.46 | 8,778 | 17.07 |
| NIPPV-S BR 10/10 | 252 | 98.10 | 4,058 | 21.19 |
| NIPPV-S BR 15/10 | 306 | 97.16 | 10,016 | 20.92 |
| NIPPV-S BR 20/10 | 368 | 97.63 | 11,511 | 19.95 |
| NIPPV-S BR 25/10 | 376 | 98.26 | 9,827 | 15.59 |
| Mean | 297.8 | 97.52 | 8,838 | 18.94 |
| NIPPV-D BR 5/5 | 230 | 95.07 | 9,694 | 12.45 |
| NIPPV-D BR 10/10 | 313 | 94.01 | 20,314 | 7.79 |
| NIPPV-D BR 15/10 | 385 | 96.02 | 19,928 | 9.68 |
| NIPPV-D BR 20/10 | 554 | 94.43 | 37,903 | 6.39 |
| NIPPV-D BR 25/10 | 875 | 94.55 | 74,115 | 6.45 |
| Mean | 471.4 | 94.82 | 32,391 | 8.55 |
| Mean for HFNC, NIPPV-S, NIPPV-D combined | 372 | 96.12 | 18,167 | 13.53 |
| NIPPV-S EXE 20/10 | 2,115 | 95.42 | 134,194 | 5.72 |
| NIPPV-D EXE 20/10 | 3,119 | 92.39 | 374,125 | 5.46 |
| Mean | 2617 | 93.9 | 254,160 | 5.59 |
| HFNO Talk 60 | 5,358 | 97.68 | 144,974 | 21.49 |
| HFNO EXE 60 | 3,495 | 94.52 | 245,883 | 6.45 |
| HFNO FEV 60 (n=10) | 276,682 | 97.94 | 1,981,211 | 16.34 |
| *HFNO FEV 60 (n=9)* | *36,175* | *95.31* | *348,345* | *9.91* |
| HFNO Cough 60 | 40,505 | 99.23 | 402,607 | 22.44 |

| | | | | |
|---|---|---|---|---|
| Mean | 72443 | 96.93 | 624,604 | 15.32 |
| Breath, mask | 120 | 96.98 | 5,955 | 15.46 |
| Talk, mask | 2,770 | 98.56 | 63,056 | 22.44 |
| Exercise, mask | 3,581 | 94.17 | 272,758 | 7.50 |
| Shout, mask | 10,202 | 98.62 | 180,760 | 18.31 |
| FEV, mask | 5,119 | 92.51 | 541,382 | 3.28 |
| Cough, mask | 13,174 | 97.04 | 313,536 | 5.59 |
| Mean * | 6969 | 96.18 | 274,298 | 11.42 |
| Mean across all activities and procedures* | 21894 | 96.43 | 226,833 | 13.99 |

Table S6: The total average numbers of particles per person sampled in 1 minute, and their estimated total volume, with the percent of each of these contributed by particles ≤5 µm during six different activities, with and without non-invasive ventilation, and with masks. Both FEV manoeuvrers and cough were performed six times in the sample. BR is breath, EXE is exercise, HFNO is high flow nasal oxygen with flows in L/min, NIPPV is non-invasive positive pressure ventilation, either single circuit (S) or dual circuit (D), with the IPAP/EPAP pressures in cm $H_2O$. HFNO FEV 60 is shown twice, for all 10 subjects, and with n=9, after removal of an extreme outlier.

* Excluding quiet breathing

#### Table S7 Fold Difference in Particle Counts when Compared to Breathing Alone across all Subjects and when the Highest is Removed.

| Fold Change Compared to Breath | All Subjects (N=10) | Highest Removed (N=9) |
|---|---|---|
| Talk | 34.63 | 31.12 |
| Exercise | 58.01 | 55.77 |
| Shout | 163.64 | 147.26 |
| FEV | 227.58 | 164.69 |
| Cough | 370.76 | 323.81 |
| HFNC BR 20 | 1.27 | 1.33 |
| HFNC BR 40 | 1.73 | 1.67 |
| HFNC BR 60 | 2.30 | 1.94 |
| HFNO TLK 60 | 32.26 | 27.35 |
| HFNO EXE 60 | 39.49 | 36.97 |
| HFNO FEV 60 | 156.58 | 90.14 |
| HFNO CHG 60 | 182.80 | 174.18 |
| NIPPV-D BR 5/5 | 1.86 | 2.00 |
| NIPPV-D BR 10/10 | 2.87 | 2.64 |
| NIPPV-D BR 15/10 | 3.06 | 3.04 |
| NIPPV-D BR 20/10 | 4.57 | 4.71 |
| NIPPV-D BR 25/10 | 7.77 | 7.01 |
| NIPPV-S BR 5/5 | 1.49 | 1.31 |
| NIPPV-S BR 10/10 | 1.35 | 1.37 |
| NIPPV-S BR 15/10 | 2.14 | 2.15 |
| NIPPV-S BR 20/10 | 2.43 | 2.26 |
| NIPPV-S BR 25/10 | 2.64 | 2.37 |
| NIPPV-D EXE 20/10 | 39.47 | 40.23 |
| NIPPV-S EXE 20/10 | 20.64 | 20.36 |

Table S11: Comparison of fold changes for respiratory activities and NIVs all compared to breath, for all subjects (N=10) and with the subject contributing highest emission is removed (N=9). HFN is high flow nasal canula with flow in L/min, NIPPV is non-invasive ventilation (single and dual limb circuit) with IPAP/EPAP pressures shown in cm $H_2O$. BR refers to breathing, TLK, talk; FEV, forced expiratory volume; CHG, cough; EXE, exercise.

#### Table S8 Recorded Ventilator Parameters for Varying Inspiratory Pressures in Two Subject Post-Hoc Study

| Ventilator Parameter | Pressures (IPAP/EPAP) | C6 Hamilton Ventilator Mean (SD) | V60 Phillips Ventilator Mean (SD) |
|---|---|---|---|
| Tidal Volume (Litres) | 5/5 | 1.5 (0.5) | 1.2 (0.1) |
| | 10/10 | 1.6 (0.5) | 1.5 (0.2 |
| | 15/10 | 2.0 (0.4) | 1.7 (0.2) |
| | 20/10 | 2.4 (0.4 | 1.7 (0.1) |
| | 25/10 | 2.9 (0.6) | 2.2 (0.2) |
| | 20/10 EXE | 2.8 (0.5) | 2.3 (0.5) |
| | Overall, mean (SD) | 2.2 (0.7) | 1.8 (0.5) |
| | Difference in mean (%) | 24.6 (p<0.001) | |
| Respiratory Rate (BPM) | 5/5 | 16 (3.5) | 13.0 (1.8) |
| | 10/10 | 15.3 (3.6) | 12.2 (1.5) |
| | 15/10 | 14.2 (2.3) | 13.0 (5.7) |
| | 20/10 | 17.2 (5.8) | 14.7 (2.3) |
| | 25/10 | 13.7 (1.5) | 10.7 (3.8) |
| | 20/10 EXE | 16.5 (1.4) | 18.8 (2.9) |
| | Overall, mean (SD) | 15.5 (3.4) | 13.7 (4.0) |
| | Difference in mean (%) | 13.1 (p=0.03) | |
| Peak Inspiratory (Flow L/min) | 5/5 | 73.7 (14.3) | 52.4 (5.8) |
| | 10/10 | 82.9 (31.8) | 73.2 (4.6) |
| | 15/10 | 94.8 (28.9) | 113.6 (10.2) |
| | 20/10 | 133.2 (33.2) | 147.1 (10.4) |
| | 25/10 | 174.3 (17.8) | 175.4 (9.4) |
| | 20/10 EXE | 179.7 (8.9) | 187.7 (26.0) |
| | Overall, mean (SD) | 123.1 (48.6) | 124.9 (52.2) |
| | Difference in mean (%) | -1.4 (p=0.71) | |
| Peak Expiratory (Flow L/min) | 5/5 | 34.4 (10.8) | 33.4 (5.4) |
| | 10/10 | 31.6 (5.7) | 40.5 (8.6) |
| | 15/10 | 40.3 (7.6) | 50.1 (11.5) |
| | 20/10 | 59.1 (10.4) | 60.3 (10.7) |
| | 25/10 | 67.4 (13.6) | 73.0 (16.2) |
| | 20/10 EXE | 99.7 (16.8) | 101.0 (18.5) |
| | Overall, mean (SD) | 55.4 (26.2) | 59.7 (25.6) |
| | Difference in mean (%) | -7.2 (p=0.04) | |
| Minute Volume (L/min) | 5/5 | 23.2 (4.4) | 15.4 (0.9) |
| | 10/10 | 23.6 (6.6) | 18.1 (3.2) |
| | 15/10 | 28.6 (7.5) | 22.0 (8.8) |
| | 20/10 | 39.7 (8.4) | 25.5 (3.3) |
| | 25/10 | 38.9 (5.9) | 23.1 (9.3) |
| | 20/10 EXE | 46.0 (9.6) | 43.1 (3.1) |

| | | | |
|---|---|---|---|
| | Overall, mean (SD) | 33.3 (11.1) | 24.5 (10.5) |
| | Difference in mean (%) | 36.0 (p<0.001) | |
| MV Leak [‡] (L/min) | 5/5 | 1.8 (0.7) | |
| | 10/10 | 2.2 (0.7) | |
| | 15/10 | 1.6 (1.8) | |
| | 20/10 | 3.3 (1.4) | |
| | 25/10 | 2.2 (2.0) | |
| | 20/10 EXE | 1.0 (0.3) | |
| | Overall, mean (SD) | 2.0 (1.4) | |
| Total Leak [§] (L/min) | 5/5 | | 20.2 (3.8) |
| | 10/10 | | 25.5 (2.4) |
| | 15/10 | | 24.0 (2.5) |
| | 20/10 | | 24.0 (6.2) |
| | 25/10 | | 26.5 (7.7) |
| | 20/10 EXE | | 30.5 (9.1) |
| | Overall, mean (SD) | | 25.1 (6.3) |
| ASI (%) [†] | Overall, mean (SD) | 29.8 (18.9) | 1.0 (2.4) |

Table S7: Data is presented as a combined mean for both subjects for each different pressure, including an overall mean across all pressures for each ventilator. Below this is a percentage difference between the overall means for each ventilator across all pressures for each parameter. P-values were obtained from a paired T-test. During exercise (EXE), subjects received non-invasive ventilation at pressures of 20/10cmH20 after achieving 70% of estimated maximal heart rate via peddling on an exercise bike for the duration of the recording. IPAP denotes, Inspiratory Positive Airway Pressure; EPAP, Expiratory Positive Airway Pressure; EXE, Exercise; BPM, Breaths per minute; L/min, Litres per minute; ASI, Asynchrony Index.

†ASI is presented as a percentage. The following algorithm was used to calculate ASI: (asynchronous events ÷ total respiratory rate) ×100. Flow waveforms analysed by investigators at time of recording. ASI expressed as a mean across all pressures excluding exercise.

‡Derived by delivered volume divided by exhaled volume. Represents leak at the patient/mask interface as calculated by the ventilator in litres per minute.

§Total Leak detected in ventilator system in litres per minute.

### References.

1. Kenward MG, Roger JH. Small Sample Inference for Fixed Effects from Restricted Maximum Likelihood. Biometrics 1997;53(3):983.
2. Ingebrethsen BJ, Cole SK, Alderman SL. Electronic cigarette aerosol particle size distribution measurements. Inhal Toxicol [Internet] 2012 [cited 2021 Jan 11];24(14):976–84. Available from: https://pubmed.ncbi.nlm.nih.gov/23216158/
3. Hess DR. Patient-ventilator interaction during noninvasive ventilation. Respir Care [Internet] 2011 [cited 2020 Nov 2];56(2):153–67. Available from: https://pubmed.ncbi.nlm.nih.gov/21333176/
4. Thille AW, Rodriguez P, Cabello B, Lellouche F, Brochard L. Patient-ventilator asynchrony during assisted mechanical ventilation. Intensive Care Med [Internet] 2006 [cited 2021 Jan 11];32(10):1515–22. Available from: https://pubmed.ncbi.nlm.nih.gov/16896854/
5. Hamilton. Hamilton-C6 Operators Manual [Internet]. 2017. [cited 2021 Jan 11];Available from: https://www.hamilton-medical.com/dam/jcr:32ec370d-3b9e-45b4-92d4-0f40fef81bd3/HAMILTON-C6-OpsMan-v1.x.x-EN-624945.01.pdf
6. How to improve patient-ventilator synchrony. Waveform analysis and optimization of ventilator settings - HealthManagement.org [Internet]. [cited 2021 Jan 11];Available from: https://healthmanagement.org/c/icu/whitepaper/how-to-improve-patient-ventilator-synchrony
7. Respironics. Respironics V60/V60 Plus Ventilator User Manual.
8. Vasconcelos RDS, Melo LHDP, Sales RP, et al. Effect of an automatic triggering and cycling system on comfort and patient-ventilator synchrony during pressure support ventilation. Respiration [Internet] 2014 [cited 2021 Jan 11];86(6):497–503. Available from: https://pubmed.ncbi.nlm.nih.gov/24051384/
9. Optiflow fitting instructions [Internet]. [cited 2021 Jan 11];Available from: https://resources.fphcare.com/content/fitting-guide-optiflow-nasal-cannula-fitting-guide-ui-620171.pdf
10. Fisher M. RT045 XS/S/M/L Non-Vented Hospital Full Face Mask with Anti-Asphyxiation Valve Single Use. 2019.
11. Royal Australian College of General Practioners. Putting on a Mask [Internet]. 2020. [cited 2021 Jan 11];Available from: https://www.racgp.org.au/FSDEDEV/media/documents/Running a practice/Practice resources/Putting-on-a-mask.PDF
12. Aerotrak. 9500 optical particle counter manual [Internet]. [cited 2021 Jan 11];Available from: https://tsi.com/getmedia/8890a386-fc04-4afa-a774-a653ba32d02a/AeroTrak_Portable_9500_A4_5001279?ext=.pdf
13. Morawska L, Johnson GR, Ristovski ZD, et al. Size distribution and sites of origin of droplets expelled from the human respiratory tract during expiratory activities. J Aerosol Sci 2009;40(3):256–69.
14. Chao CYH, Wan MP, Morawska L, et al. Characterization of expiration air jets and droplet size distributions immediately at the mouth opening. J Aerosol Sci [Internet] 2009 [cited 2021 Jan 11];40(2):122–33. Available from: https://pubmed.ncbi.nlm.nih.gov/32287373/
15. Nicas M, Nazaroff WW, Hubbard A. Toward Understanding the Risk of Secondary Airborne Infection: Emission of Respirable Pathogens. J Occup Environ Hyg [Internet]

2005 [cited 2021 Jan 16];2(3):143–54. Available from: https://www.tandfonline.com/action/journalInformation?journalCode=uoeh20
16. Stadnytskyi V, Bax CE, Bax A, Anfinrud P. The airborne lifetime of small speech droplets and their potential importance in SARS-CoV-2 transmission. Proc Natl Acad Sci U S A [Internet] 2020 [cited 2021 Jan 16];117(22):11875–7. Available from: https://doi.org/10.5281/zenodo.3770559.
17. Eaton W, Bax A, Netz R. Physics of Virus Transmission by Speaking Droplets. Proc Natl Acad Sci [Internet] 2020 [cited 2021 Jan 16];2020.05.12.20099630. Available from: https://doi.org/10.1101/2020.05.12.20099630
